# Machine learning-driven COVID-19 early triage and large-scale testing strategies based on the 2021 Costa Rican Actualidades survey

**DOI:** 10.1101/2024.04.02.24305223

**Authors:** Carlos Pasquier, Maikol Solís, Vivian Vilchez, Santiago Núñez-Corrales

## Abstract

The COVID-19 pandemic underscored the importance of mass testing in mitigating the spread of the virus. This study presents mass testing strategies developed through machine learning models, which predict the risk of COVID-19 contagion based on health determinants. Using the data from the 2021 “Actualidades” survey in Costa Rica, we trained models to classify individuals by contagion risk. After theorize four possible strategies, we evaluated these using Monte Carlo simulations, analyzing the distribution functions for the number of tests, positive cases detected, tests per person, and total costs. Additionally, we introduced the metrics, *efficiency* and *stock capacity*, to assess the performance of different strategies. Our classifier achieved an AUC-ROC of 0.80 and an AUC-PR of 0.59, considering a disease prevalence of 0.26. The fourth strategy, which integrates RT-qPCR, antigen, and RT-LAMP tests, emerged as a cost-effective approach for mass testing, offering insights into scalable and adaptable testing mechanisms for pandemic response.

## 1 Introduction

During the SARS-Cov-2 pandemic, local and global authorities underscored the importance of mass testing as a non-pharmacological measure to control the spread of the disease. In this article, we propose population-level strategies of disease mass testing that leverage mechanisms for predicting individual contagion risk. We hypothesize that the infection status depends on a prior set of individual and collective factors (Kelly, 2021; Shahbazi et al., 2020; Gesesew et al., 2021; Abate et al., 2020).

Approximately between 5 and 10 days is the latent period between the infection/incubation, and spreading phases for COVID-19 (Tindale et al., 2020). Any mass testing strategies must consider the presence of pre-symptomatic and asymptomatic individuals (Oran and Topol, 2021, 2020; Slifka and Gao, 2020). It includes additional factors must be considered, such as clinical history, population behavior, and geography, transforming the optimization problem into a non-linear resource allocation problem with memory (Kırkızlar et al., 2010; Du et al., 2021). Therefore, the efforts to control the spread of the disease must focus on detecting and isolating these populations to effectively curb disease spread before solutions like vaccines become available.

In the shortage of resources during the pandemic, health institutions required reliable testing at scale technologies (Mercer and Salit, 2021). The gold standard for COVID-19 testing, RT-qPCR, proved the most reliable with its high sensitivity and specificity (Yang and Rothman, 2004). However, clinical centers reliance on specialized laboratories and trained staff, as well as reagent availability, hampers its global applicationWiencek et al. (2020). Antigen-based tests offer a less expensive, rapid alternative but suffer from lower sensitivity (Mercer and Salit, 2021; Peeling et al., 2021; CDC, 2020). Another molecular testing technology, RT-LAMP, provides sensitivity and specificity comparable to RT-qPCR, with lower biosafety requirements and faster results Mautner et al. (2020); Amaral et al. (2021).

In Costa Rica, COVID-19 testing predominantly relied on RT-qPCR, despite its limitations and high costs Segura-Ulate et al. (2022). The 7-day average positivity rate reached up to 60% in September 2020, with testing capacity remaining constant until December 2020 (Núñez-Corrales and de Camino, 2021). In September 2020, regulations for antigen-based testing were introduced (MINSA, 2021b), but their use only began in December 2021. Despite increased RT-qPCR testing capacity in the private sector, the positivity rate remained above 10%. Costa Rica did not implement a mass testing strategy against COVID-19, even though it could help control the pandemic at a lower societal and economic cost compared to mobility restrictions.

A more balanced approach partitions the population into high-risk and low-risk groups based on their exposure risk, tailoring testing strategies accordingly. The high-risk group includes symptomatic individuals, healthcare workers, and essential workers (Haley et al., 2023). RT-qPCR or similar technologies like RT-LAMP testing is crucial for this group to prevent superspreading events. The low-risk group consists of individuals with limited exposure, such as those who telecommute or practice social distancing. This group is the target for mass testing campaigns to capture all the events in batches (Millioni and Mortarino, 2021). We propose two strategies for the low-risk group: a pooling technique and a multiple testing scheme. The pooling technique divides the total number of individuals into different pools, testing each group to maximize the number of tests with reduced time, money, and chemical reagent costs (Millioni and Mortarino, 2021). The multiple testing scheme involves weekly or biweekly tests, which has been shown to reduce positivity rates and sick leaves (Plantes et al., 2021; Haigh and Gandhi, 2021; Larremore et al., 2021).

The partition of the high and low risk group are defined through a machine learning algorithms (Escobar et al., 2022; Park et al., 2022). The classifier allows optimizing each strategy for each group reducing the overall number of tests applied and costs. The machine learning will use the data from the *“Actualidades 2021”* survey (Escuela de Estadística, 2021), which has socioeconomic and demographic information of a sample of people living in Costa Rica in the month of October 2021. Also, it has evaluations about the self-infections, perception of the disease and other broader topics.

This paper is organized as follows. Section 2 describes the determinants of health and the conceptual framework for testing strategies. Section 3 describes the data collection process, data wrangling, and the machine learning models used to predict COVID-19 contagion risk. Section 4 presents the results of the machine learning models and the proposed mass testing strategies. Finally, Section 5 presents the conclusions and future work.

## 2 Related work

### 2.1 Determinants of health

The World Health Organization (WHO) conceptual framework for health determinants guides the selection of variables used in this study (World Health Organization, 2010). The framework broadly categorizes health determinants into two groups: structural determinants and intermediate determinants. Structural determinants encompass the social, economic, and environmental conditions that influence health, while intermediate determinants directly affect health outcomes.

The circumstances in which individuals are born, live, and grow, known as the determinants of health, can result in health inequalities and inequities, leading to vulnerable populations or groups at risk of poor physical, psychological, or social health. Vulnerable populations often face greater health risks and disparities, especially during pandemics like COVID-19. The COVID-19 pandemic highlighted the significant challenges and disruptions faced by vulnerable populations. Social determinants of health, such as poverty, unemployment, poor housing conditions, and lack of access to healthcare services, exacerbated the impact of the pandemic, resulting in increased risks of disease, mortality, and long-term health consequences (Lima-Serrano, 2022).

From the literature, we can identify the following determinants of health that are relevant to the spread and severity of COVID-19:

#### Socioeconomic Factors

Ethnicity, low socioeconomic status, low educational attainment, low income, and unemployment have been associated with higher risks of COVID-19 infection, hospitalization, and mortality (Oppenheimer-Lewin et al., 2022; Abrams et al., 2022). Poverty is strongly linked to COVID-19 risk and adverse outcomes, compounded by life stage, poor housing conditions, and lack of access to healthcare (Abrams et al., 2022).

#### Age and Resilience

Older adults have been disproportionately affected by COVID-19, experiencing higher mortality rates (Oppenheimer-Lewin et al., 2022). However, they may experience less psychological distress compared to other groups due to resilience and effective coping strategies (Oppenheimer-Lewin et al., 2022). Resilience, influenced by social determinants of health, plays a vital role in adapting to adversity during the pandemic (Oppenheimer-Lewin et al., 2022).

#### Mental Health

Mental health disorders increase vulnerability to COVID-19, exacerbating symptom severity and leading to increased suicide rates during the pandemic (Lima-Serrano, 2022). High loneliness, social isolation, and depressive symptoms are associated with lower resilience and risk factors for chronic diseases (Oppenheimer-Lewin et al., 2022).

#### Housing and Homelessness

Overcrowded housing, poor building conditions, lack of access to clean water and sanitation, and inadequate infrastructure contribute to the spread and severity of COVID-19 (Galanis and Hanieh, 2021). Homeless populations face challenges in accessing healthcare, hygiene supplies, and vaccination services (Abrams et al., 2022).

#### Food Insecurity and School Closures

Food insecurity increased during the pandemic, affecting access to balanced meals, especially among children (Abrams et al., 2022). School closures exacerbated this issue, and food delivery measures were implemented to address the need (Abrams et al., 2022). Furthermore, school closures had a detrimental impact on educational achievement, particularly for children facing adverse social determinants (Abrams et al., 2022).

Other hidden factors can contribute as well to the infection and spreading of Sars-Cov-2. In the next sections we will describe the determinants of health defined in our dataset.

### 2.2 Conceptual framework for testing strategies

Let us first define some common notations used across the whole study. Denote as *D*_*P*_ be the condition of having the disease (i.e. infected) and *D*_*N*_ the condition of being no infected. The prevalence is estimated by ℙ (*D*_*P*_) such that ℙ (*D*_*N*_) = 1 − ℙ (*D*_*P*_). Let also *N* be the total population to undergo testing. Thus, *N × ℙ* (*D*_*P*_) are the true infected and *N ×* (1− *ℙ* (*D*_*P*_)) the true healthy people. Denote as 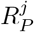 and 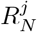 the results positive and negative of each test, respectively. In addition, let *j* = PCR, Ag or LAMP denote each available testing technology, RT-qPCR, Antigen or RT-LAMP respectively. We can thus define sensitivity the proportion of people infected who are correctly identified as positive in the test, or 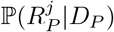. Specificity constitutes the proportion of people not infected who are correctly identified as negative in the test, or 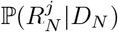.

According to the literature, we use RT-qPCR (sensitivity ≥90%, specificity ≥90%), Antigen (sensitivity ≈80%, specificity≥90%), and RT-LAMP (sensitivity ≥90%, specificity ≥95%).

We can improve the mass testing effectiveness by using techniques like pooling or multiple testings Pooling strategies, like the one-dimensional (1D) protocol, involve mixing samples taken in batches and analyzing them together (Dorfman, 1943). This method is useful only at low prevalence levels and may suffer from limitations such as loss of sensitivity due to dilution or sample collection artifacts Millioni and Mortarino (2021); Mercer and Salit (2021). Multiple testing, on the other hand, offers an alternative when pooling is infeasible. The optimal testing frequency and isolation periods are influenced by factors like transmission rates and disease prevalence, which are often unpredictable Du et al. (2021); Sandmann et al. (2020).

During the pandemic, the Costa Rican Ministry of Health followed CDC recommendations and defined guidelines for antigen-based testing as an alternative to RT-qPCR in both public and private health services (MINSA, 2021b; CDC, 2020). A key distinction between the two systems is the requirement of a confirmatory RT-qPCR test for negative antigen-based results in the public sector, while the private sector is exempt from this. High-risk patients are classified based on the presence of symptoms and epidemiological nexus, and are directed to the public healthcare system, while low-risk patients are directed to the private system.

A successful mass testing strategy should rapidly screen individuals while minimizing false negatives and positives. We propose a two-step strategy that first classifies patients into high and low risk categories, and then applies a suitable adaptive mass testing strategy per group. The strategy relies on patient data to predict risk categories, with symptomatic patients or those with an epidemiological nexus automatically classified as high-risk. Based on the predicted risk category, different testing strategies are applied to high and low-risk groups. The effectiveness of each strategy depends on the prevalence, sensitivity, specificity, positive predictive value (PPV), and negative predictive value (NPV) of each test, as well as the accuracy of the predictive model.

## 3 Methods

This section describes the data collection process, data wrangling, and the machine learning models used to predict COVID-19 contagion risk. We also describe the proposed mass testing strategies and the simulation framework used to evaluate their effectiveness.

### 3.1 Dataset

The Department of Statistics in the Universidad de Costa Rica, in collaboration with senior students, conducted a national survey on COVID-19 and other related socioeconomic and opinion in Costa Rica. They performed the survey on October 2021. They included a probabilistic sample of 2003 adults who are mobile phone users. Due to the ongoing COVID-19 pandemic, traditional face-to-face interviews were not feasible. Therefore, the survey relied on institutional resources, such as random digit dialing, a random mobile phone number generator, and the use of 60 VPNs (Virtual Private Networks) to enable IP (Internet Protocol) telephony for personnel to conduct interviews from their personal computers at home. The survey was designed in Spanish and took approximately 15 minutes to complete.

The survey collected information to risk exposure towards COVID-19, diverse opinions about the pandemic, and socioeconomic information. The survey design, refinement, interview execution, coding and tabulation took four-month to complete.

### 3.2 Data Wrangling

For processing the data in the survey, we used the pandas library in Python. The original set consisted of 129 columns with 2003 observations. We removed columns that were not relevant to the analysis such as date or survey identification. We removed variables related opinions about the country situation, hypothetical situations or possible biases around mental health and COVID-19. Due to the relevance of the study, we prefer to focus on a actual risk factors related to the disease.

We prefixed all the variables with sd_ or id_ representing the structural and intermediate determinants of the health respectively. Then, after the prefix we added a code representing the determinant of health. Table 1 shows all the coded variables.

**Table 1:** Prefixes for determinants of health used in the variables.

| Structural | Intermediate |
| --- | --- |
| `sd_edu`: Education | `id_beh`: Behavioral |
| `sd_eco`: Economic | `id_bio`: Biological |
| `sd_occ`: Occupation | `id_psy`: Psychosocial |
| `sd_inc`: Income | `id_mat`: Material |
| `sd_eth`: Ethnicity |  |
| `sd_gen`: Gender |  |
| `sd_cul`: Cultural |  |

In the survey, the variable rp_1 was used to identify if the respondent had COVID-19. We recorded into the new variable covid19 as True, if rp_1 is *“Si, lo tuvo”* (*“Yes, I had it”*), *“Si, lo tiene”* (*“Yes, I have it”*). Otherwise, False if rp_1 is *“No”*. The variable collects the disease condition given by the respondent, and it is not a medical

diagnosis.

The next step was to clean inconsistencies, re-leveling and re-coding variables. To re-level the variables we use 2 (High risk), 1 (Moderate risk) and 0 (Low risk). Binary variables were re-coded as 1 (True) in the presence of risk or condition and 0 (False) otherwise. The pre-processing merged almost all variables due to they presented questions around a specific topic. In those cases, we used the rounded median of the values to create a new variable. After this, we finished with 29 variables related with the health and environmental status, socioeconomic status, behavioral attitude towards the pandemic and cultural aspects like religion. The pre-processing pipeline imputed the missing variables with the mean for the numeric variables: weight (15.38%), height (6.64%), member in a household with 18+ years old (3.2%) and total members in the household (4.24%). Only two categorical needed imputation using the most frequent value: Perception of contagion (3.25%) and anxiety symptoms (0.25%). The complete set of variables and their meaning used in the study are in the supplementary Appendix in Table A1.

### 3.3 Machine learning models

We used AutoML library from H2O (LeDell and Poirier, 2020), to automatically adjust the classification model. The hyperparameters pipeline tuning used Random grid search. We split the dataset into 80% for training and 20% for testing stratified by the variable covid19. The pipeline included Distributed Random Forest (including Extremely Randomized Trees), Generalized Linear Models, XGBoost, Gradient Boosting Machines and Stacked Ensembles^1^. We excluded Deep Learning models because the relative small size of the dataset, and the high computational cost of the training process for little gains in the accuracy.

The pipeline generated, optimized and tested the mentioned models for predicting the condition of having COVID-19, the remaining variables are the features (Table A1). It used a five-fold cross-validation, followed by a testing with held-out dataset. These variables have 1464 negatives and 539 positives indicating a 26.90% of prevalence. Given the imbalanced proportion of positive, we used the Precision-Recall area under the curve (AUC-PR) as criteria to select the best model. The AUC-PR is a metric that combines the precision and recall of a model, and it is more robust on these cases.

### 3.4 A Proposal for Mass Testing Strategy

Following the technical details from Solís et al. (2022) we denote a high-risk classification outcome by *H*, and a low-risk one by *L*. The model’s sensitivity ℙ (*H*|*R*_*P*_) and specificity ℙ (*L*|*R*_*N*_)

In the context of antigen-based testing, we denote as *S*_−5_ the event of a patient having less than 5 days since symptom onset and _+5_ otherwise. We assume that *S*_−5_ and *S*_+5_ are independent of high-risk conditions or test results. In our computational experiments, we set ℙ (*S*_+5_) with values of 25%, 50%, and 75%. We define ℙ (*S*_−5_) = 1− ℙ (*S*_+5_). A greater proportion of RT-qPCR tests are used directly on high-risk patients when P(*S*_+5_) increases, and the number of antigen-based tests used at the group level increases when *P* (*S*_−5_) increases.

Table 2 summarizes the four strategies considered in this study. The first strategy is a baseline used by The Costa Rica government during the pandemic. The second strategy is using a pooling technique over the low risk population to maximize coverage and reduce costs. The third strategy is a consecutive antigen testing, which was a common practice in the private sector. The fourth strategy is a saliva testing with RT-LAMP, which is a promising alternative to RT-qPCR.

**Table 2:** Strategies settings in the simulation framework.

|  | Risk level |  |
| --- | --- | --- |
| | Low ($L$) | High ($H$) |
| **Strategy 1:**<br>**antigen-based testing** | No testing | • Patients in $S_{-5}$ are tested with an Antigen test.<br>– Positive results $R_P^{Ag}\vert H$ go to quarantine.<br>– Negative results $R_N^{Ag}\vert H$ are tested with a RT-qPCR test.<br>• Patients in $S_{+5}$ are tested with a RT-qPCR test. |
| **Strategy 2:**<br>**pooling** | Dorfman pooling technique with 5 samples. Both negatives and positives are declared as is. | Same as Strategy 1. |
| **Strategy 3:**<br>**consecutive antigen testing** | All patients are tested with an Antigen test:<br>• Negative results $R_N^{Ag}\vert L$ are accepted as is.<br>• Positive results $R_P^{Ag}\vert L$ are retested again within one or two weeks. | Same as Strategy 1. |
| | Low ($L$) | High ($H$) |
| **Strategy 4:**<br>saliva testing using RT-LAMP | Same as Strategy 3. | Split the High risk group into: Regular High risk ($H$) and the Essential workers ($E$) (medics, police, etc.).<br>• The Essential workers all are tested with RT-qPCR<br>• All the regular High risk group is tested with RT-LAMP:<br>• Positive results $R_P^{LAMP}\vert H$ go to quarantine.<br>• Negative results $R_N^{LAMP}\vert H$ need a confirmatory with RT-LAMP. |

### 3.5 Mathematical formulation for massive testing strategy simulation

The first element to consider is the number of high-risk and low-risk patients, *N*_*H*_ and *N*_*L*_ respectively. We assume that the number of patients is a random variable with a Binomial distribution. Thus, we can define:

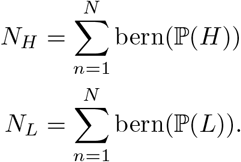

where bern(*p*) represents the outcome of Bernoulli random variable with probability *p*. We will denote the number of tests applied for each technology as 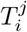, where *j* stands for the testing technology to use or any indication about the variable, and *i* = 1, 2, 3, 4 stands for the strategy. The same notation applies for the number of positive cases reported, 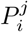, and the cost, 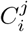.

### 3.6 Number of tests

For the number of tests for each strategy we can estimate the following:

***Strategy 1***

In the high-risk group, we need to separate the number of Antigen and RT-qPCR test given they belongs to *S*_−5_ or *S*_+5_.

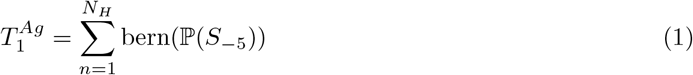

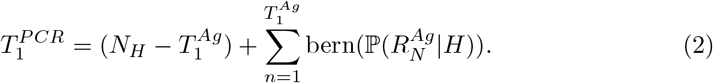

***Strategy 2***

Antigen-based tests remain unchanged at 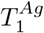 as Equation (1). We divide the RT-qPCR tests in this strategy into 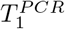 as Equation (2) and tests for the pooling strategy.

The prevalence in the low-risk group is the false omission rate *p*_*L*_ = 1 − *ℙ* (*R*_*N*_ | *L*). With an assumed constant sensitivity of RT-qPCR tests 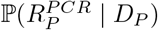, the number of positive groups is calculated as:

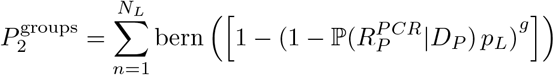

The total number of tests required for pooling is:

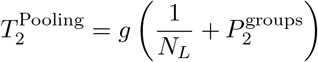

Thus, the overall number of RT-qPCR tests is:

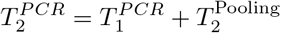

### Strategy 3

The antigen-based test count, considering re-testing, has two distinct parts. In the case of the high-risk group, we adopt the Strategy 1, denoted as 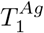. Conversely, for the low-risk group, an initial test is applied to all. Those testing positive are then subjected to a second test. Consequently, the aggregate antigen-based test count for re-testing is represented by:

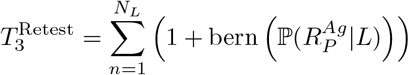

Here, 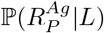 is the probability of being positive given a member of the low-risk group. The total count for antigen-based tests is obtained as:

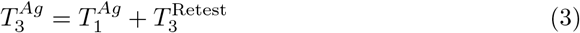

### Strategy 4

In this strategy we introduce a new testing technology, RT-LAMP, which is applied to the essential workers like doctors, nurses, police officers, etc.

Define the number of Essential workers as *N*_*E*_ as

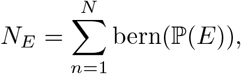

where ℙ (*E*) is the probability of being an essential worker. For simulation purposes, we set as 1%. The value is a conservative estimate based on the 1.25% of total healthcare workers in Costa Rica: 2470 in the Ministry of Health, 62814 in the public social security from a total population of 5213374 inhabitants (MINSA, 2021a; CCSS, 2021; Brenes Camacho et al., 2013). Therefore we define

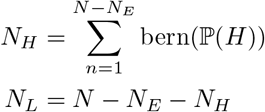

The number of Antigen-based tests are estimated with Equation (3) as Strategy 3.

The other estimates per technology are:

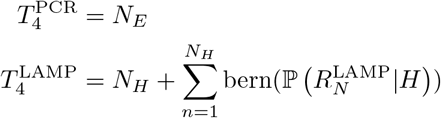

### 3.7 Positive detection

The second key element is the number of positive cases detected by each strategy. As before we split the models per strategy and risk level.

#### Strategy 1

We simulate the number of negative individuals with antigen-based given they were classified as high-risk as

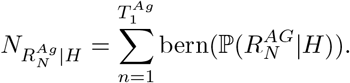

The number of cases reported is given by:

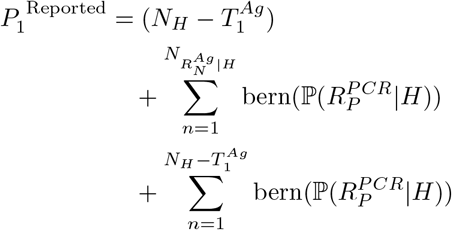

#### Strategy 2

According to Dorfman (1943), the probability of having a positive test overall into the groups are:

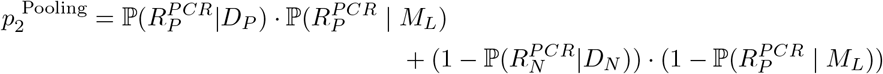

where we denoted 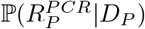 as the sensitivity and 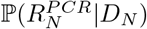 as the specificity

of the RT-qPCR test respectively.

For the number of positive cases reported, we have again two components. First, we have the same number as Strategy 1 for the high-risk population. For the low-risk branch, we need to consider only those groups with positive test outcomes. We estimate the probability that their individual test in the Dorfman scheme attains a positive result,

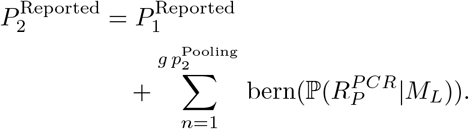

#### Strategy 3

First, we have the same number of positive cases as Strategy 1 for the high-risk population. For the low-risk branch, we need to consider only the test that were positives in the first or second round. This estimate is defined as

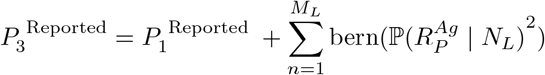

#### Strategy 4

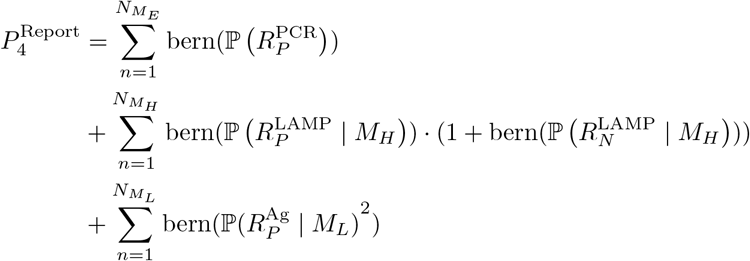

### 3.8 Total cost, tests per person and performance across strategies

The total costs for each strategy are computed as the sum of the costs for each technology. Table 3 summarizes the formulas used in each case

**Table 3:** Costs.

| Strategy | Cost | Test per person |
| --- | --- | --- |
| 1 | $C_1 = C^{\text{Ag}} T_1^{\text{Ag}} + C^{\text{PCR}} T_1^{\text{PCR}}$ | $T_1^{\text{per person}} = \frac{T_1^{\text{Ag}} + T_1^{\text{PCR}}}{N_H}$ |
| 2 | $C_2 = C^{\text{Ag}} T_2^{\text{Ag}} + C^{\text{PCR}} T_2^{\text{PCR}}$ | $T_2^{\text{per person}} = \frac{T_2^{\text{Ag}} + T_2^{\text{PCR}}}{N}$ |
| 3 | $C_3 = C^{\text{Ag}} T_3^{\text{Ag}} + C^{\text{PCR}} T_1^{\text{PCR}}$ | $T_3^{\text{per person}} = \frac{T_3^{\text{Ag}} + T_1^{\text{PCR}}}{N}$ |
| 4 | $C_4 = C^{\text{PCR}} T_4^{\text{PCR}} + C^{\text{LAMP}} T_4^{\text{LAMP}} + C^{\text{Ag}} T_4^{\text{Ag}}$ | $T_4^{\text{per person}} = \frac{T_4^{\text{Ag}} + T_4^{\text{PCR}} + T_4^{\text{LAMP}}}{N}$ |

### 3.9 Strategies assumptions

We define the cost of a single RT-qPCR test as *C*^*P CR*^ = $100 and the cost of an antigen-based test as *C*^*Ag*^ = $50, assuming a total population of *N* = 1000 individuals. We use the logit transformation to estimate the probabilities of being classified as high or low risk depending on the testing technology and the classifier’s sensitivity and specificity. The overall cost, number of tests per person, and number of positive reported cases for each strategy are defined in the following sections. The proposed mass testing strategy provides a framework for informed decision-making in the context of COVID-19 testing and can be adapted based on the specific characteristics of the population and testing technologies available.

In the next sections we present the results for the costs, number of positive cases and number of tests per person for each strategy. We also show two measures called *stock capacity* and *efficiency*. The stock capacity is the number of tests that can be bought per dollar. The efficiency is the number of positive cases detected per dollar spent. We refer to the reader to Solís et al. (2022) for all the technical details.

## 4 Results

### 4.1 Model’s metrics and results

We present in Table 4, the summary statistics of the processed data.

**Table 4:**
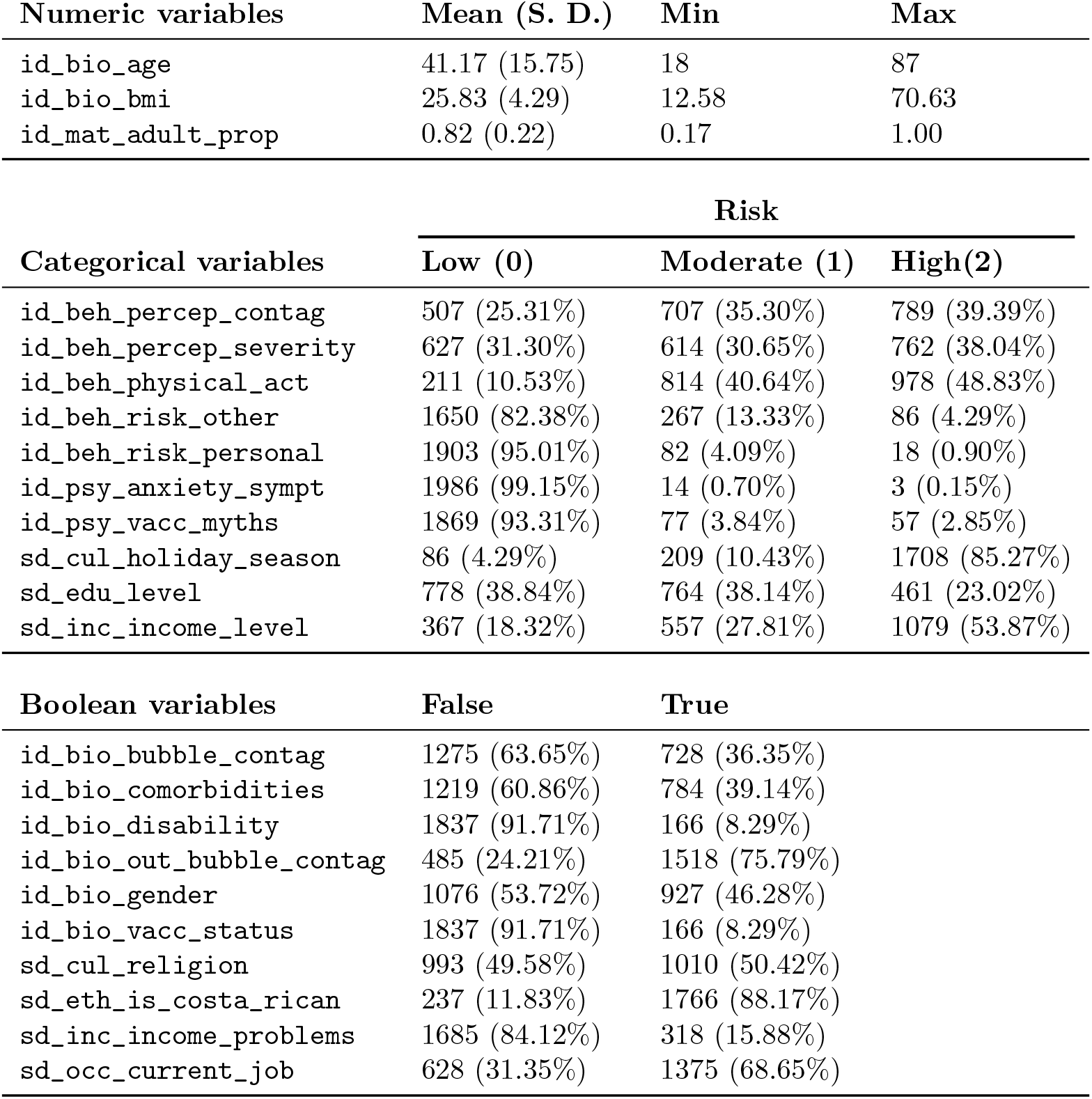
Summary statistics.

After this feature engineering, classification models like logistic regression, Random Forest, Gradient Boosting and XGBoost were adjusted with their respective hyperparameters. Using these models, we obtained in the test sample under a decision threshold of 0.3037 (defined based on the maximization of the F1 metric). Taking this threshold into account, we estimate some metrics of interest in Table 5. Also, the confusion matrix is shown in Table 6.

**Table 5:**
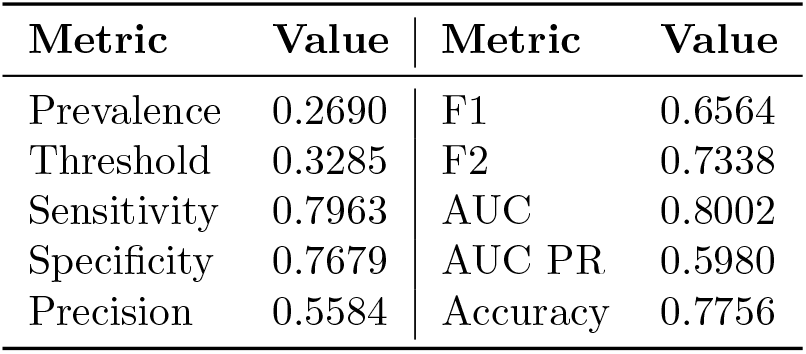
Selected machine learning model summary metrics.

| **Metric** | **Value** | **Metric** | **Value** |
| --- | --- | --- | --- |
| Prevalence | 0.2690 | F1 | 0.6564 |
| Threshold | 0.3285 | F2 | 0.7338 |
| Sensitivity | 0.7963 | AUC | 0.8002 |
| Specificity | 0.7679 | AUC PR | 0.5980 |
| Precision | 0.5584 | Accuracy | 0.7756 |

**Table 6:** Confusion matrix (Act/Pred) with a threshold of 0.3037.

|  |  | **Predicted** |  | **Error Rate** |
| --- | --- | --- | --- | --- |
|  |  | **False** | **True** |  |
| **Actual** | **False** | 225 | 68 | 0.2321 |
|  | **True** | 22 | 86 | 0.2037 |
| **Total** |  | 247 | 154 | 0.2244 |

Figure 1 shows the receiver operating characteristic (ROC) and precision-recall (PR) curves for the model. The ROC curve plots the false positive rate (1 - specificity) against the true positive rate (sensitivity or recall). The PR curve plots the recall against the precision. Both use different thresholds to calculate the metrics. The area under the of curve (AUC) of the ROC curve for this model is 80.02% indicates a better classification than random. The area under the PR curve (AUC-PR) compares the false positive rate is more useful than the AUC when the dataset is imbalanced, as the number of true negatives results are not used. The AUC-PR for this model is 59.80%. It means the model predicts correctly the double of positive cases against using a biased classifier with only 26.90% (prevalence) of being positive.

**Fig. 1:**
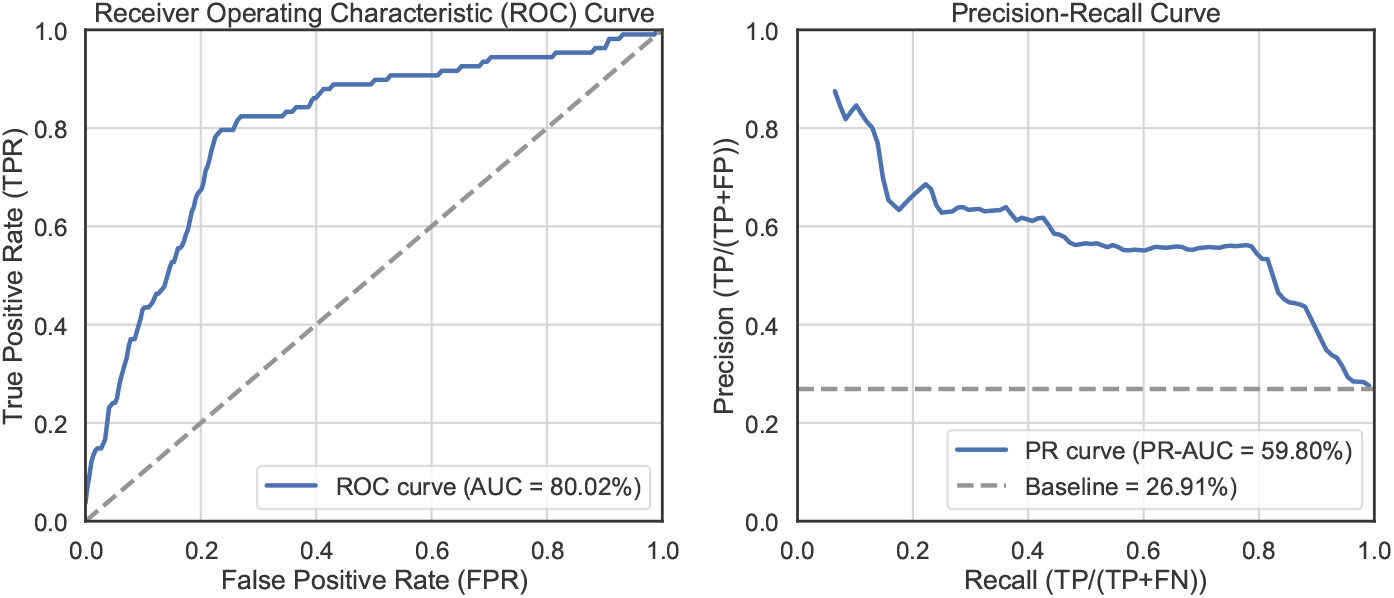
ROC and PR curves for the classification model.

To determine the most relevant variables inside the model, we quantified the SHAP (SHapeley Additive Explanation) values and variable importance analysis.

The SHAP values estimate the computing the marginal contribution of each feature to the model’s output, while holding all other features constant. This allows us to understand how each feature influences positively (red) or negatively (blue) the prediction. Figure 2 has these values for our model.

**Fig. 2:**
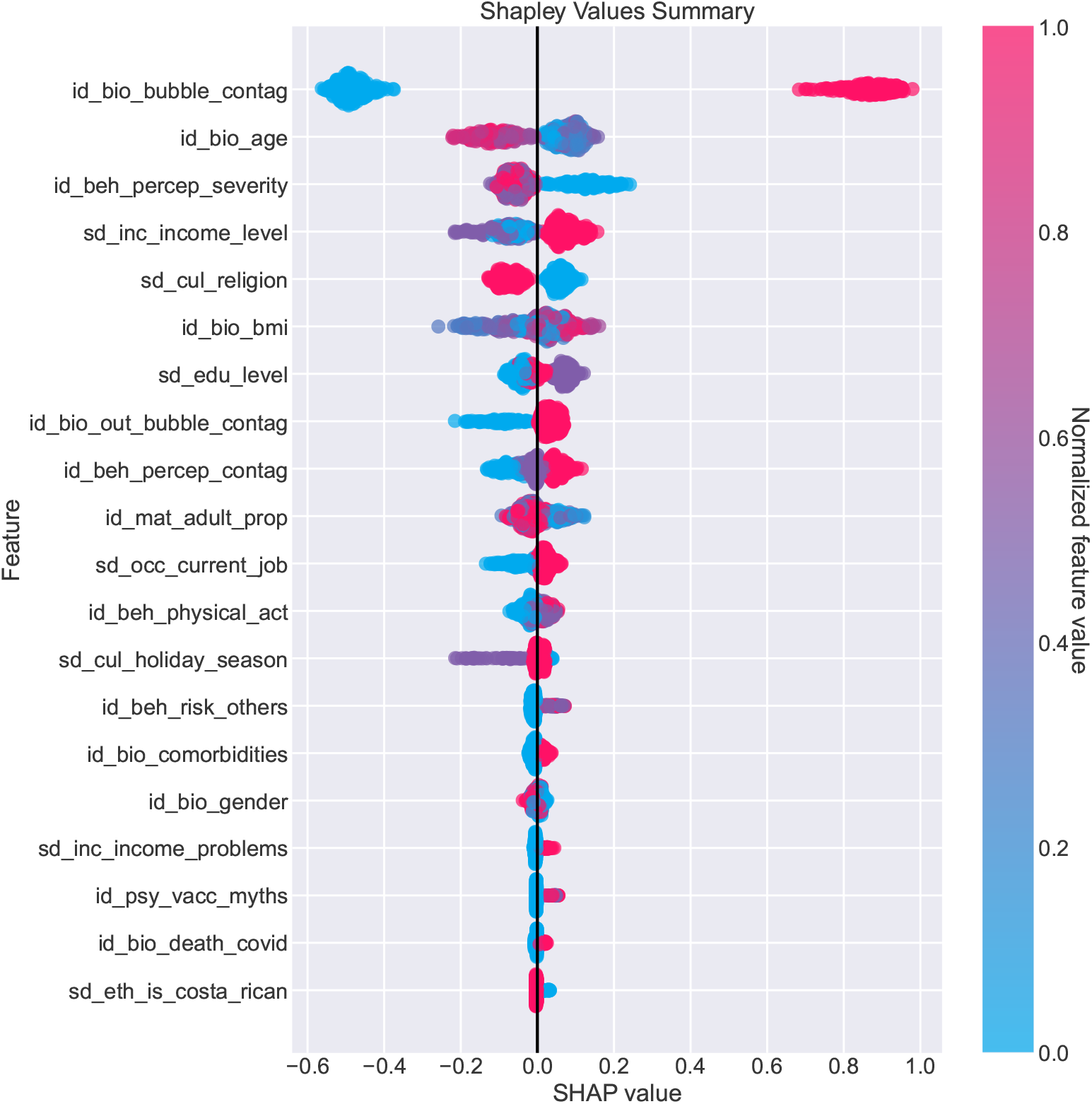
SHAP values for the classification model.

The most prominent feature on the plot is the *contagion inside the bubble*. This is expected given the multiple reports around the influence of physical proximity to spread the disease (Fazio et al., 2021). The *age* has an inverse relation where older people have fewer chances to get infected. This is a natural consequence of a relative young workforce in Costa Rica. The third most relevant variable is the *severity perception*. Here people with moderate or high scores (4 or plus) tend to have less chance of infections.

A relevant variable is the *income level*. Notice that high income individual has more chances of being infected. In a broader sense, this group has more resources to travel abroad, commute to other regions in the country or interact with other people due to work related tasks. *Religion* become a relevant feature. The informational campaigns mainly around Catholics cause a decrease of the infection chances. Other measures become informative like *BMI, education level* or *contagion outside the bubble*.

In Figure 3, we present the variable importance analysis. We opted to use the permutation variable importance method described in Fisher et al. (2019). The method permutes every variable one-at-time and estimates the difference on the prediction gains against the fitted model. Give the random nature of the procedure, we took 100 repetitions for each variable and then normalized to sum up 1. In the case the

**Fig. 3:**
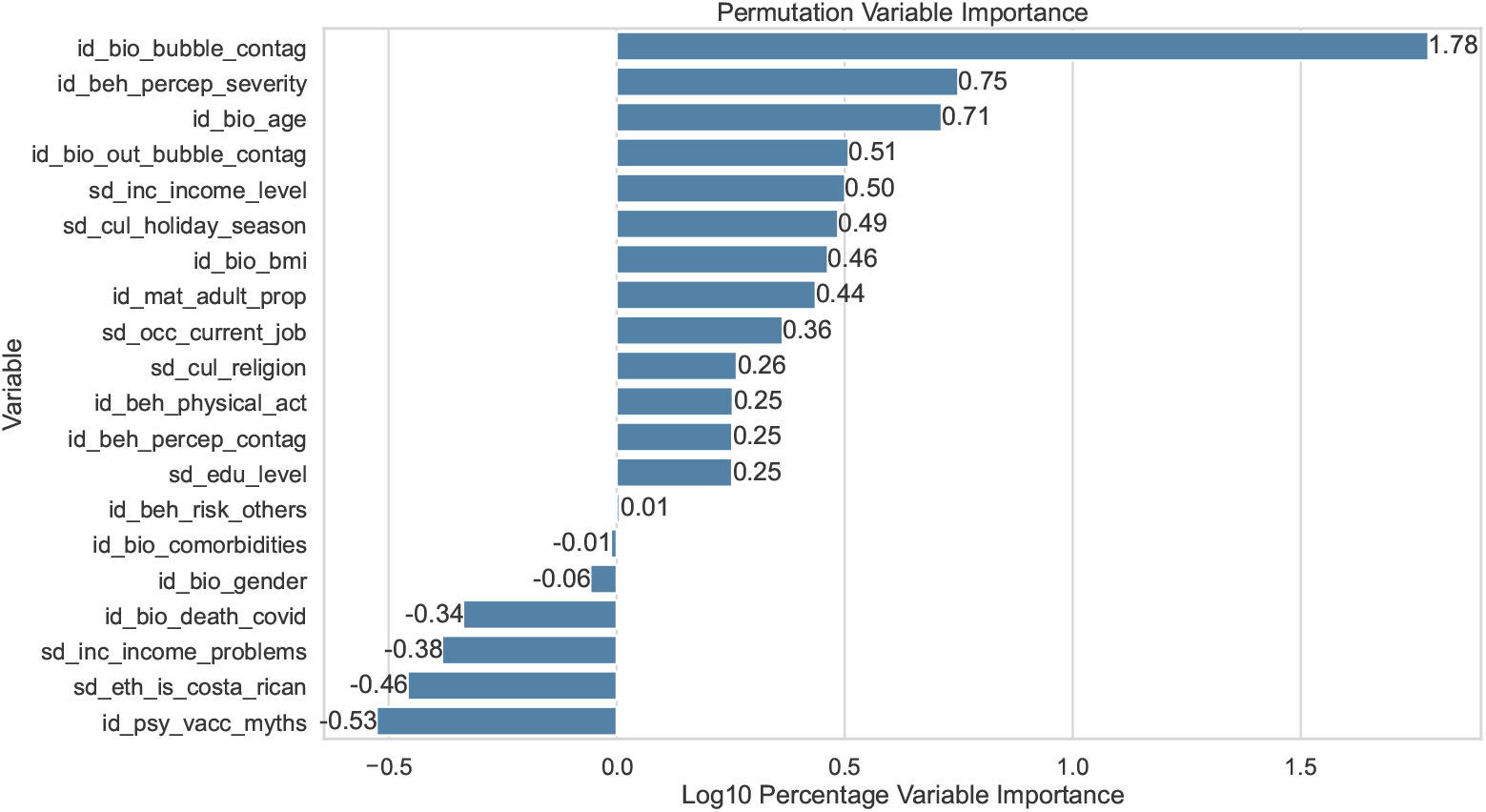
Variable importance for the classification model.

The results agree with the SHAP values, where the *contagion inside the bubble* is the most predominant feature. Then they follow *severity perception, age, contagion outside the bubble* and *income level*.

### 4.2 Monte Carlo simulations of mass testing strategies

To evaluate the economic and epidemiological impact of the strategies developed in Solís et al. (2022), we used the data that was used for training the model, as was the number of positives and negatives estimated by the model to give an approximation of the prevalence of COVID-19 for the date of the data. We also used the sensitivity and specificity of the model that was previously developed. Using Monte Carlo simulation, we simulated different epidemiological and cost scenarios (1000 simulations) using 1000 people to test the strategies.

As seen in Figure 4, the distribution of the number of positives detected tends to be higher for strategy 4, followed by strategy 2 and with similar behaviors to each other, strategies 1 and 3. This tells us, in particular, that in terms of positive people reported with COVID-19, the one with the highest number of reported people would be strategy 4, where this behavior is more evident when the proportion of people with symptoms greater than 5 days grows.

**Fig. 4:**
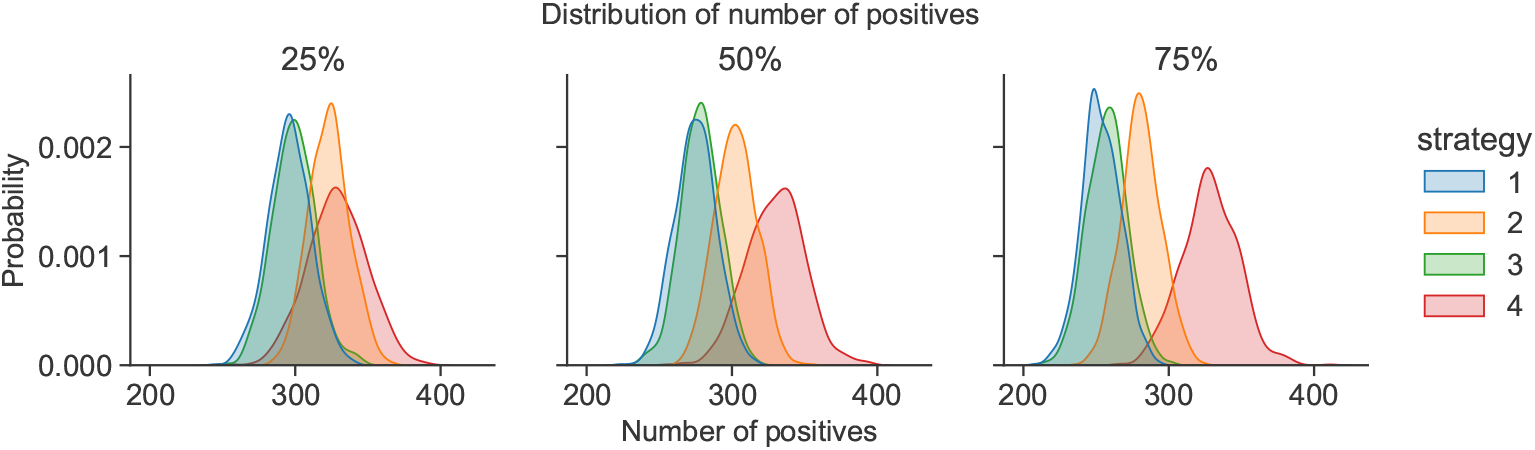
Distribution of the number of positive reports by strategies and by proportion of people with onset of symptoms greater than 5 days

Figure 5 shows that the strategy that performs the least tests per person is strategy 2 under any proportion of symptom onset greater than 5 days in the population. This is followed by strategy 3 and the order of strategy 1 and 4 depending on the proportion of occurrence of symptoms.

**Fig. 5:**
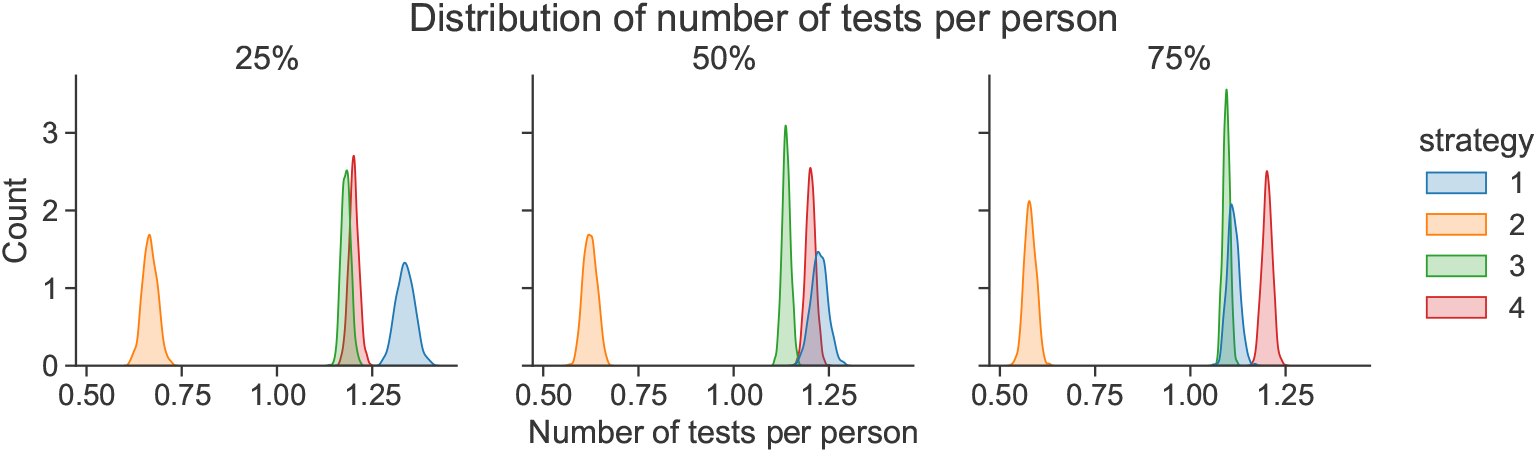
Distribution of the number of tests carried out by strategies and by proportion of people with onset of symptoms greater than 5 days

Figure 6 shows the distribution of costs for each strategy. As can be seen, the one that would have the lowest cost would be strategy 1, followed by strategies 4, 2 and 3. However, it is important to note that although one strategy is cheaper than another, its effectiveness must also be evaluated through their ability to detect positives. This can be seen better in figure 7:

**Fig. 6:**
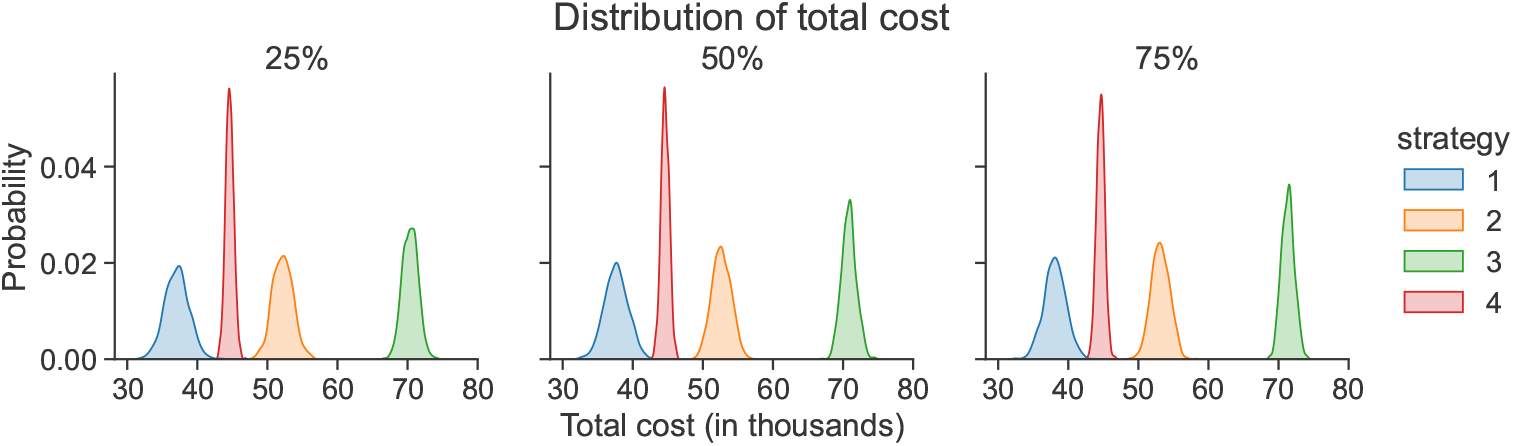
Distribution of the total cost by strategies and by proportion of people with onset of symptoms greater than 5 days

**Fig. 7:**
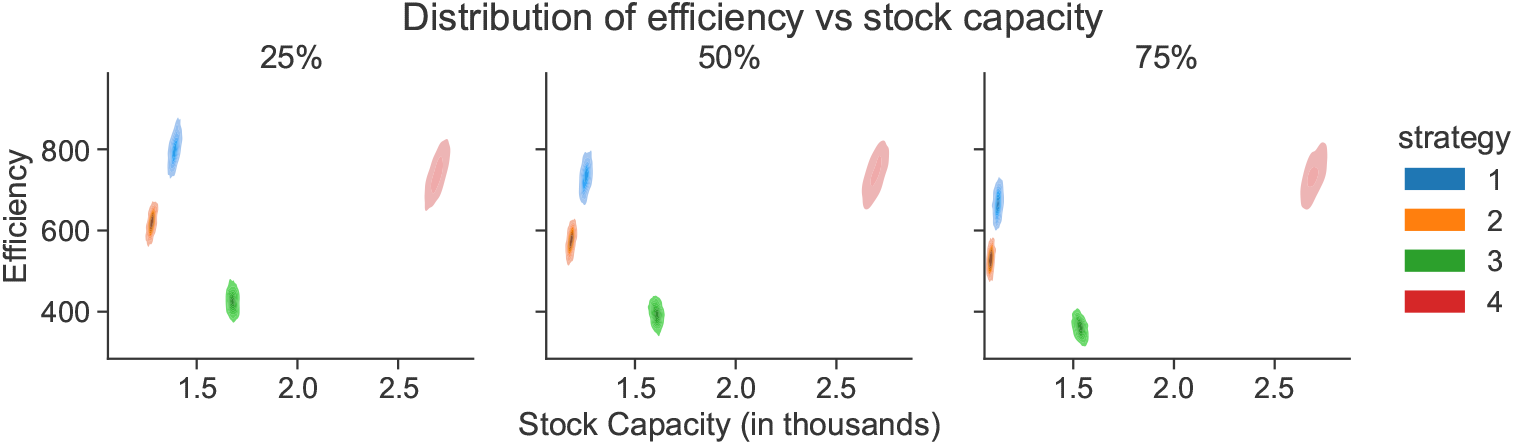
Distribution of efficiency by strategy and by proportion of people with onset of symptoms greater than 5 days

Figure 7 shows a graph of efficiency (efficiency defined as the ability of each strategy to detect one positive case per dollar spent) and stock capacity (represents the number of tests that can be purchased given a budget). As shown, the strategy with the highest efficiency and highest stock capacity is strategy 2. Strategies 1 and 4 tend to be efficient at detecting a positive case but not as efficient when considering a budget. Strategy 3 tends to be inefficient and with a not-so-high stock capacity.

## 5 Conclusions

This paper has investigated the use of mass testing strategies tailored to specific risk groups and testing technologies to improve the efficiency and impact of testing campaigns during pandemics. In the case of Costa Rica, the proposed strategies were designed according to the *MINSA* official guidelines.

We targeted the risk groups using a machine learning model trained with the determinants of health extracted from the *“Actualidades 2021”* survey. The model achieved an AUC of 0.80 and an AUC-PR of 0.59, indicating a better classification than random. The most relevant variables were *contagion inside the bubble, severity perception, age, contagion outside the bubble* and *income level*. The model can be used to predict the risk of COVID-19 infection in Costa Rica and can be adapted to other countries by using local data.

The proposed mass testing strategies were evaluated using a Monte Carlo simulation. The results showed that the strategy with the highest efficiency and highest stock capacity is Strategy 2. Strategies 1 and 4 tend to be efficient at detecting a positive case but not as efficient when considering a budget. Strategy 3 tends to be inefficient and with a not-so-high stock capacity. For Strategy 4, the inclusion of RT-LAMP as a testing technology has shown to be a promising alternative to address the limitations of antigen-based tests. RT-LAMP offers high sensitivity and specificity comparable to RT-qPCR but at a lower cost and with simpler equipment and reagents. Additionally, RT-LAMP provides results within an hour, facilitating faster decision-making and contact tracing efforts.

This approach has the potential to enhance the limited resources assigned to *MINSA* and other health institutions during pandemics. We need to work on the inclusion of other epidemiological variables like the number of cases, hospitalizations and deaths to improve the model’s performance. Furthermore, the adoption in the national guidelines of machine learning mechanism can help to handle the uncertainty and speed-up the decision during pandemics.

## Data Availability

All data produced in the present study are available upon reasonable request to the authors.

## Acknowledgements

The authors would like to thank to Vicerrectoría de Investigación and the departaments of Escuela de Matemática and Centro de Investigación en Matemática Pura y Aplicada (CIMPA) of Universidad de Costa Rica for the financial and administrative support in the development of this research through the project 821-C2198.

## Appendix A Cleaned variables

**Table A1:**
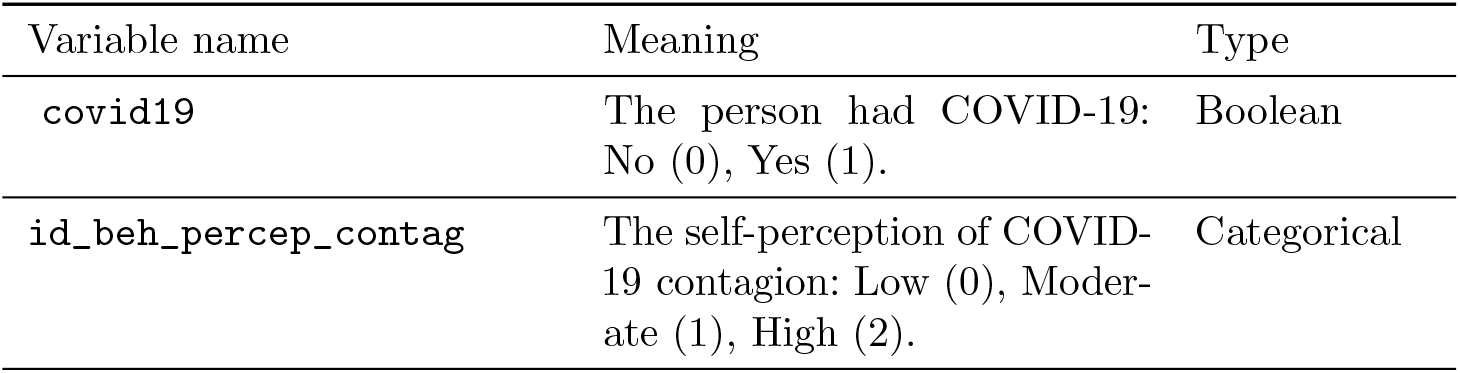
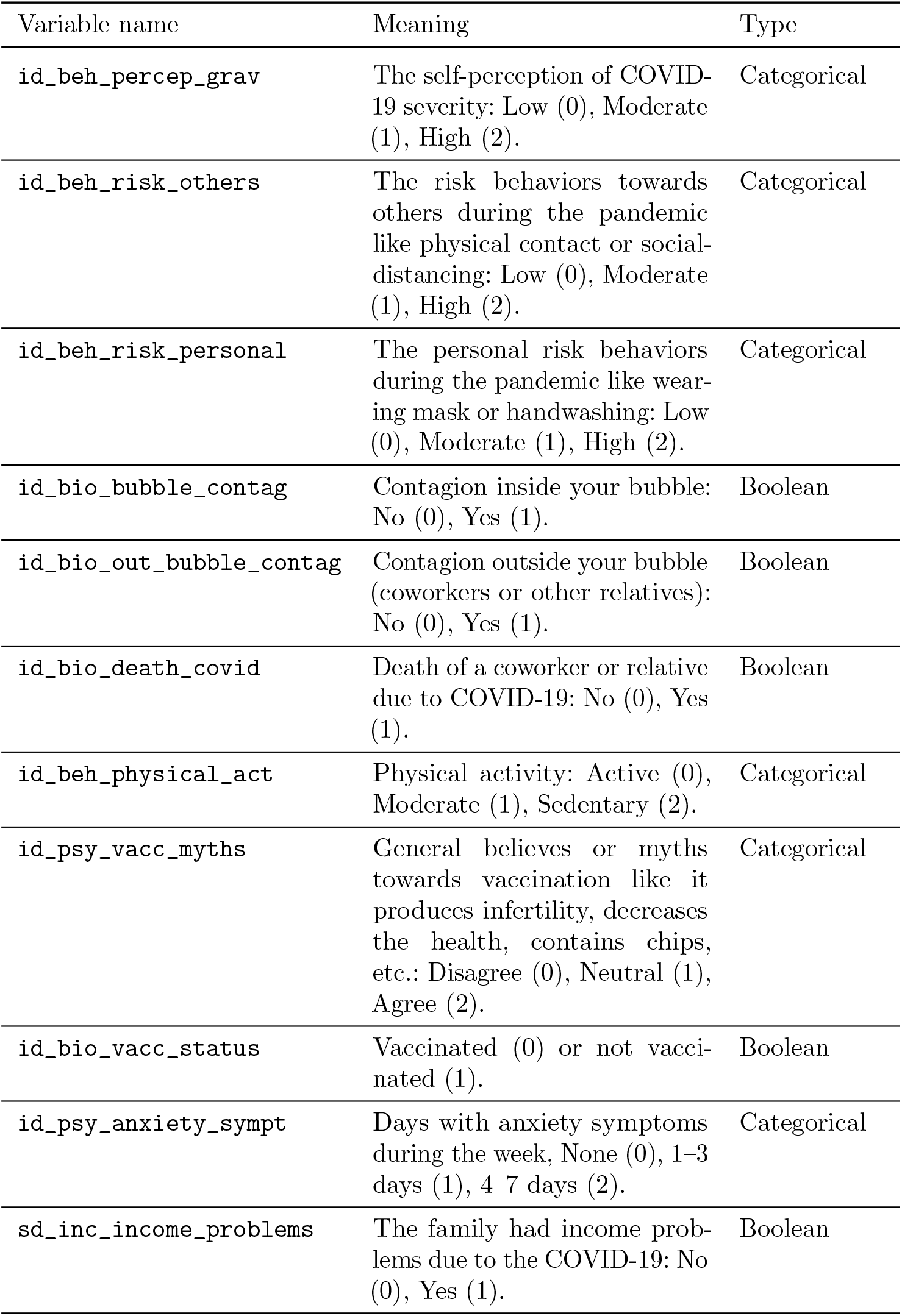
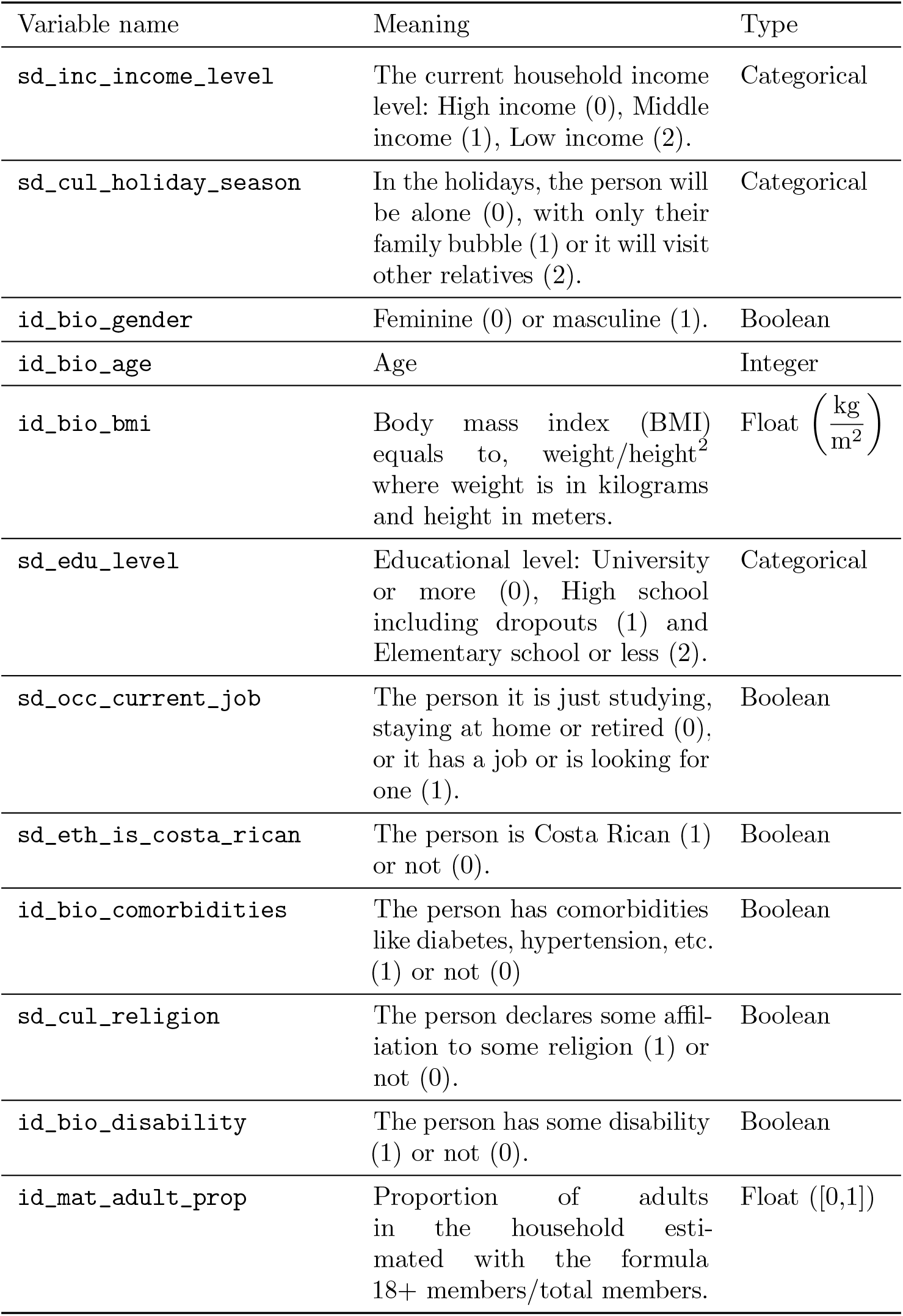
Cleaned variables. Boolean variables are 0 (False) and 1 (True), while Categorical variables are as 0 (Low Risk), 1 (Moderate Risk) and 2 (High Risk).

## Footnotes

1 The list of hyperparameter optimized for each algorithm is available in the H2O website https://docs.h2o.ai/h2o/latest-stable/h2o-docs/automl.html

